# A Bibliometric Analysis of Research Hotspots and Frontiers in Diabetes and Cardiovascular Disease Comorbidity from 2005 to 2024

**DOI:** 10.1101/2024.09.29.24314577

**Authors:** Jingjin Wang, Tianyuan Su, Yunlong Li, Yinxia Su, Yukai Li

**Author notes:** Corresponding author: Yinxia Su, 567 Shangde North Road, Shuimogou District, Urumqi City, Xinjiang 830000, China, Yukai Li, 393 Xinyi Road, Xinshi District, Urumqi City, Xinjiang 830000, China. These authors contributed euqally,and thus they were joint-first authors. Jingjin Wang, Yinxia SU and Yukai Li takes responsibility for all aspects of the reliability and freedom from bias of the data presented and their discussed interpretation. E-mail address of all authors: JW; TS; Yl; YS; YL.

## Abstract

Nearly two decades ago, diabetes mellitus (DM) and cardiovascular disease (CVD) emerged as major global health threats, with research interest steadily increasing. Despite ongoing in-depth studies in the DM+CVD field, a systematic analysis of overall development trends and future hotspots remains lacking. This study employs bibliometric methods, utilizing CiteSpace and VOSviewer software to analyze and visualize 5,505 DM+CVD-related publications from 2005 to May 2024. The findings reveal a rapid growth trend in the field, with publications peaking at 751 in 2022, and the United States and China as major contributing countries. Keyword analysis identifies research hotspots including oxidative stress, insulin resistance, and heart failure. Highly cited literature analysis shows that the relationship between DM+CVD and obesity, disease mechanisms, treatment strategies, lifestyle interventions, and novel drugs are primary research themes. Analysis of highly cited literature from the past three years indicates that the application of SGLT2 inhibitors and GLP-1 receptor agonists, the association between metabolic abnormalities and DM+CVD, precise management of heart failure and cardiomyopathy, and in-depth epidemiological studies are current and future research priorities. This study not only reveals research hotspots and development trends in the DM+CVD field but also provides important references for optimizing research resource allocation and promoting multidisciplinary collaboration.

---

Diabetes Mellitus (DM) is a group of chronic metabolic disorders characterized by hyperglycemia, primarily resulting from defects in insulin secretion or its biological action, with the potential for chronic damage and dysfunction of multiple organs^1^. DM is broadly classified into Type 1 Diabetes Mellitus (T1DM), Type 2 Diabetes Mellitus (T2DM), and Gestational Diabetes Mellitus (GDM). T1DM is an autoimmune disease, generally attributed to the abnormal activation of immune cells, such as CD8+ T and CD4+ T cells, which target and attack the body’s pancreatic β-cells^2^. T2DM, accounting for 90-95% of all DM cases, is primarily characterized by insulin resistance and relative insulin deficiency, typically associated with genetic predisposition, lifestyle factors, and environmental influences^2,3^. GDM refers to any degree of glucose intolerance first appearing or detected during pregnancy, potentially affecting maternal and fetal health4. Although GDM typically resolves postpartum, some women may continue to exhibit glucose metabolism abnormalities and face an increased risk of developing T2DM in the future^2,4^.Currently, the global prevalence of DM is increasing significantly, driven by demographic shifts and lifestyle changes. According to the International Diabetes Federation (IDF)^5^, the number of adults with DM reached 537 million in 2021, with projections estimating an increase to 783 million by 2045. This trend not only imposes a substantial economic burden but also significantly impacts societal development. In 2021, global healthcare expenditure attributed to DM was estimated at 966.6 billion USD, representing an approximate 43% increase from 20155. In the United States alone, the total cost associated with DM in 2022 was approximately 412.9 billion USD, comprising 306.6 billion USD in direct medical costs and 106.3 billion USD in indirect costs. These indirect costs encompass reduced employment rates, decreased work attendance, and premature mortality due to DM, collectively resulting in a productivity loss of 96.5 billion USD^6^.

Cardiovascular Diseases (CVD) are one of the primary causes of death among patients with DM^7,8^, while DM is also a major risk factor for CVD^9^. According to a report by the American Heart Association (AHA)^10^, at least 68% of DM patients aged 65 or older die from some form of CVD; in China, the proportion of T2DM patients who die from CVD is even higher, reaching 75%^11^. The chronic hyperglycemic state in DM patients leads to a series of pathological changes, such as endothelial cell dysfunction and vascular smooth muscle cell proliferation, which increase the risk of cardiovascular disease^7,12^. Furthermore, DM patients often present with a cluster distribution of risk factors, including hypertension, obesity, and dyslipidemia^2^, which further increases cardiovascular risk. Epidemiological studies indicate that, compared to non-DM populations, DM patients have a 5 to 15 percentage point higher risk of major cardiovascular events such as coronary heart disease^13^.

Despite the extensive research in the field of DM+CVD, systematic analyses of its research hotspots and frontiers remain relatively scarce. Given the profound impact of DM+CVD on patient health and socioeconomic conditions^14^, a comprehensive understanding of the research dynamics in this field is of paramount importance. Therefore, this study aims to conduct a systematic analysis of the DM+CVD field using bibliometric methods, employing two literature visualization software tools: CiteSpace and VOSviewer. Compared to traditional systematic reviews, bibliometrics offers the advantage of summarizing research fields from a macro and comprehensive perspective, utilizing mathematical and statistical models for quantitative and visual analysis of knowledge carriers^15,16^. The CiteSpace and VOSviewer software tools selected for this study each have their unique features: CiteSpace is renowned for its robust temporal perspective functionality and rich analytical indicators, while VOSviewer is known for its intuitive interface and high flexibility^17,18^.By combining the strengths of these two software tools, we can scientifically identify active researchers, potential collaborators, popular topics, and future research frontiers in the field. Through this approach, we aim to provide a comprehensive reference of hotspots and frontiers in the DM+CVD field for relevant researchers, while also elucidating the research trajectory and evolutionary trends within this domain.

## 1. Materials and Methods

### 1.1. Data Source

The primary data source for this study was the Web of Science Core Collection (WOSCC) database, utilizing the SCI-Expanded index. To comprehensively cover DM+CVD-related research, we established the following search criteria: the search term combination was (TS=(“diabetes” OR “diabetic”) AND TS=(“cardiovascular diseases” OR “cardiac diseases” OR “heart diseases”)). Document types were restricted to “Article” and “Review Article”, with all open access publications included. The language was limited to English, and the search timespan was set from January 1, 2005 to May 5, 2024.To explore the multifaceted impacts of DM+CVD in depth, we applied detailed restrictions to the citation topic Meso, encompassing but not limited to the following 34 fields (for a complete list, see Table S1): Diabetes, Phytochemicals, Lipids, Nutrition & Dietetics, and Cardiology - Circulation. Limiting these broad topic areas ensures that the research scope of DM+CVD is explored from multiple angles and levels. Through this multidisciplinary analytical approach, we can systematically evaluate research progress across various domains, from basic medicine to clinical practice, and from drug research to public health policy, thereby forming a comprehensive and in-depth analytical framework. Finally, using the aforementioned search method, 5505 publications were retrieved, including 3781 articles and 1724 reviews. The full records and references were imported into CiteSpace and VOSviewer software for quantitative analysis.

### 1.2. Analytical Methods

This study utilized CiteSpace 6.3R1 and VOSviewer software for the visualization analysis of selected publications. In the data preprocessing phase, we imported bibliographic data into CiteSpace for duplicate screening, which yielded no duplicate entries. For CiteSpace parameter configuration, the time span was set from 2005 to 2024, with annual time slices. Term sources encompassed Title, Abstract, Author Keywords (DE), and Keywords Plus (ID) to comprehensively capture key information from the literature. We employed pathfinder and pruning sliced networks algorithms for network pruning to optimize visualization outcomes. The Selection Criteria was set to TOPN, utilizing the software’s default threshold of 50, while maintaining other parameters at their default values. In VOSviewer, we flexibly adjusted thresholds for each node based on visualization requirements, with other parameters adhering to default settings.For analytical dimensions, we selected authors, publishing institutions, countries/regions, keywords, and references as primary analytical foci to comprehensively grasp collaboration patterns and knowledge structures within the field. In terms of visualization mapping, we generated collaboration network diagrams, co-occurrence network diagrams of cited literature, and co-citation network diagrams of cited literature using CiteSpace and VOSviewer to visually represent their characteristics and relationships. Additionally, Scimago Graphica was employed in conjunction with VOSviewer to provide a more intuitive display of collaboration networks and research impact across countries/regions.To elucidate core research themes and their evolution, we utilized CiteSpace’s keyword clustering functionality to conduct cluster analysis on key concepts in the research field, identifying primary research themes and corresponding concept clusters. Subsequently, we generated a Timeline-based Theme River diagram to visualize the developmental trajectories and historical evolution of different clusters. Furthermore, we constructed keyword density maps using VOSviewer to identify research hotspots and reveal the field’s overall knowledge structure.To explore research frontiers and emerging trends, we analyzed highly cited literature and their keywords from the past three years using CiteSpace. Through keyword analysis functions, particularly the Spotlight algorithm, we uncovered the strength of associations between keywords. Concurrently, by employing the Burst Detection algorithm for in-depth analysis of highly cited literature from the past three years, we successfully captured rapidly emerging research hotspots.Moreover, we utilized Excel for processing and organizing raw data, and R (version 4.3.1) with ggplot2 and forecast packages, along with Excel, to generate supplementary statistical charts and graphs.

## 2. Analysis of Bibliometric Characteristics

### 2.1 Publication Trends and Impact Analysis

From January 2005 to December 2023, the WOSCC indexed a total of 5404 articles in the field of DM+CVD, as illustrated in Figure 1. The number of publications related to DM+CVD research has demonstrated a consistent annual increase, with particularly significant increases observed during the period of 2019-2022. During this time, the annual publication volume increased substantially compared to the previous year, with an average growth rate of 21.85%. The peak in publication output was reached in 2022, with 751 articles published. Although the data for 2023 indicates a slight decline, considering the overall trend and historical data, this may be interpreted as a short-term fluctuation based on the overall trend. It is noteworthy that at the time of this study’s completion, publications for 2024 were still being continuously indexed.Given the non-linear growth characteristics of historical publication numbers, this study employed polynomial regression (second-order) to forecast the publication volume for 2024. Based on the predicted publication volume, it is estimated that 2024 will see approximately 812 publications, 95% CI [679, 946]. Judging from both the publication volume and short-term forecast results, it is evident that the attention and activity in the DM+CVD field continue to increase, garnering widespread interest from scholars.

**FIG 1.**
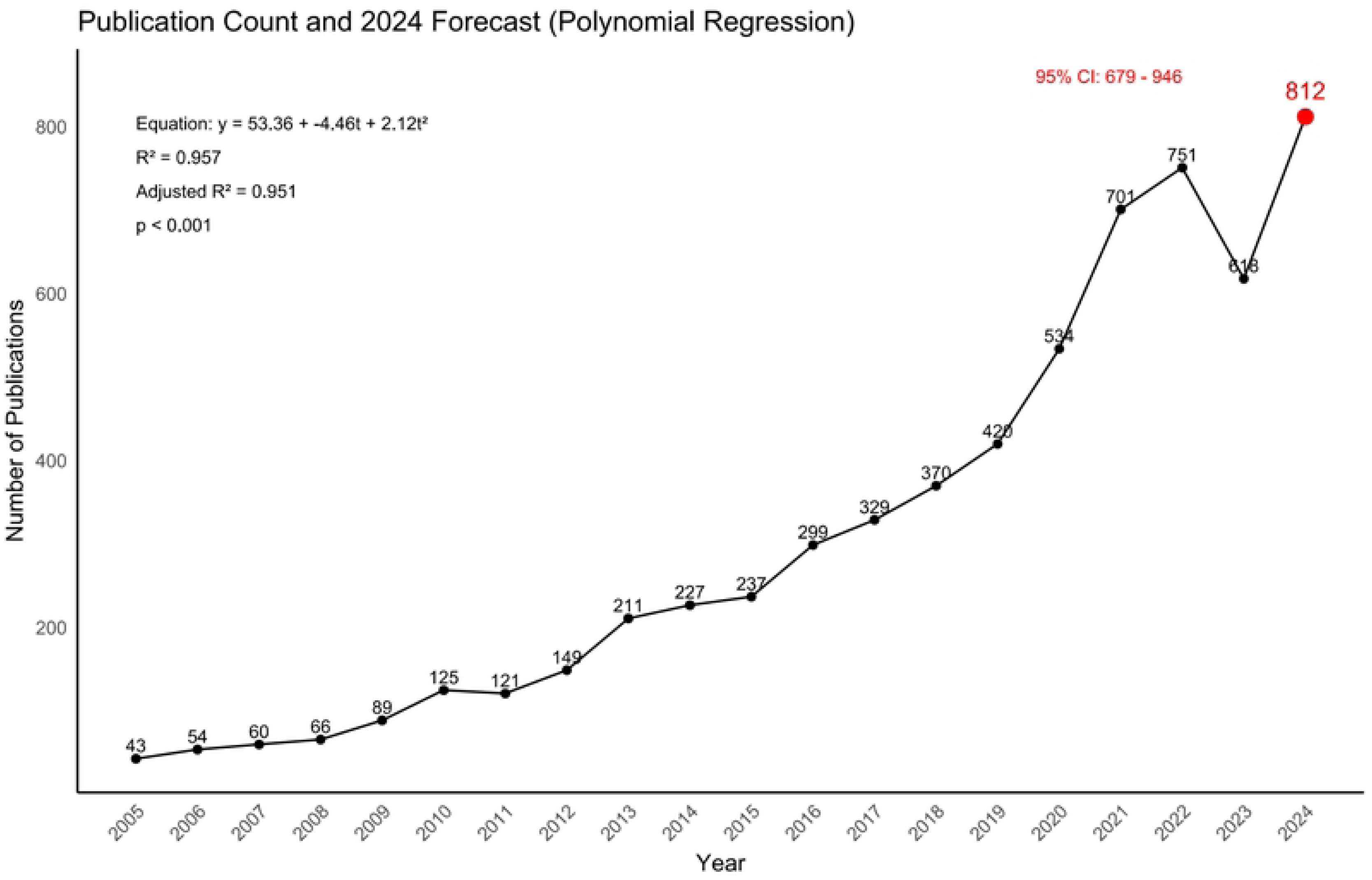
Annual publication volume in DM+CVD (2005-2024). The graph shows the annual number of publications in the field of diabetes mellitus and cardiovascular disease comorbidity (DM+CVD) from 2005 to 2023, with a forecast for 2024. Black points represent actual publication counts, while the black line shows the trend. The red point for 2024 indicates the forecasted value (812) based on polynomial regression. Equation of the regression line and statistical parameters (R², Adjusted R², p-value) are shown in the top left. The 95% confidence interval for the 2024 forecast (679-946) is displayed in the top right. Note: 2023 data may be incomplete due to ongoing indexing.

The WOS citation report function indicates that the retrieved publications have accumulated a total of 272,346 citations, with an average of 49.47 citations per article. The H-index, which reflects the impact and reputation of relevant research, stands at 212 for publications in the DM+CVD field, demonstrating the significant academic influence and the depth and durability of research outcomes in this domain.The WOS journal analysis report function reveals that all publications from 2005 to May 2024 originated from 118 journals. This study primarily analyzed the top 10 journals by publication volume, as shown in Table 1. Among these, PLoS ONE published the highest number of articles at 199, closely followed by the *International Journal of Molecular Sciences* with 159 publications. *Cardiovascular Diabetology*, *Nutrients*, and *Circulation* also maintained publication volumes exceeding 100 articles each. Notably, among the top 10 journals by publication volume, MDPI-affiliated journals account for over 30% of the publications, while BMC-affiliated journals follow closely, comprising 19% of the total.The Impact Factor (IF) is a crucial metric for assessing a journal’s academic influence. Among the top 10 journals by publication volume, *Circulation* has the highest IF at 37.8, while *Frontiers in Cardiovascular Medicine* has the lowest at 3.6. The distribution of H-indices follows a similar pattern to IF, with *Circulation* having the highest H-index of 674 and *Frontiers in Cardiovascular Medicine* the lowest at 69.In summary, research in the DM+CVD field demonstrates a sustained growth trend and exerts a profound and significant influence in academia. The continued attention from high-quality journals and high citation rates reflect the importance and research value of this field.

**TABLE 1.**
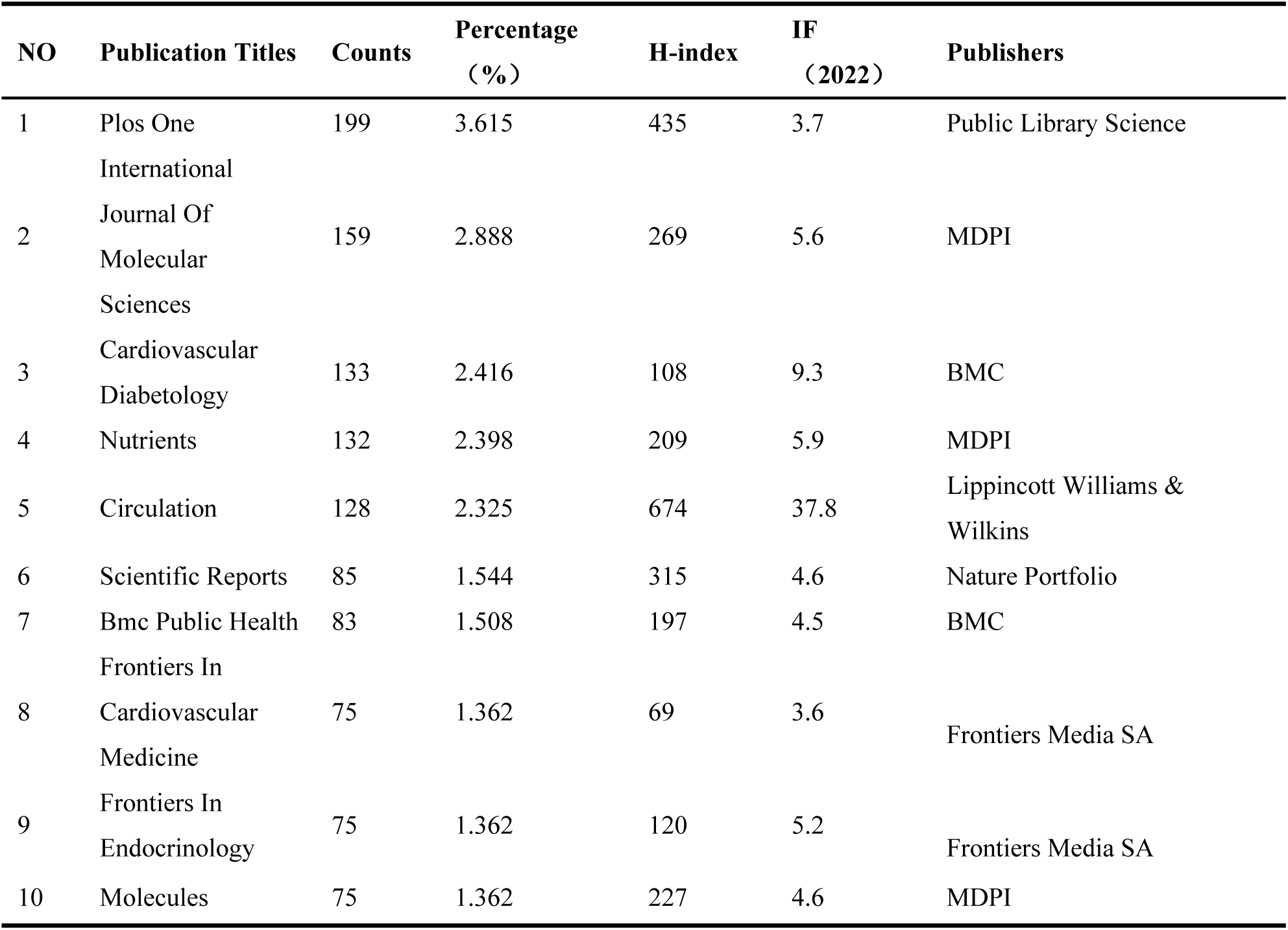
TOP 10 jorunals with publication volume from 2005 to May 2024.

**TABLE 2.**
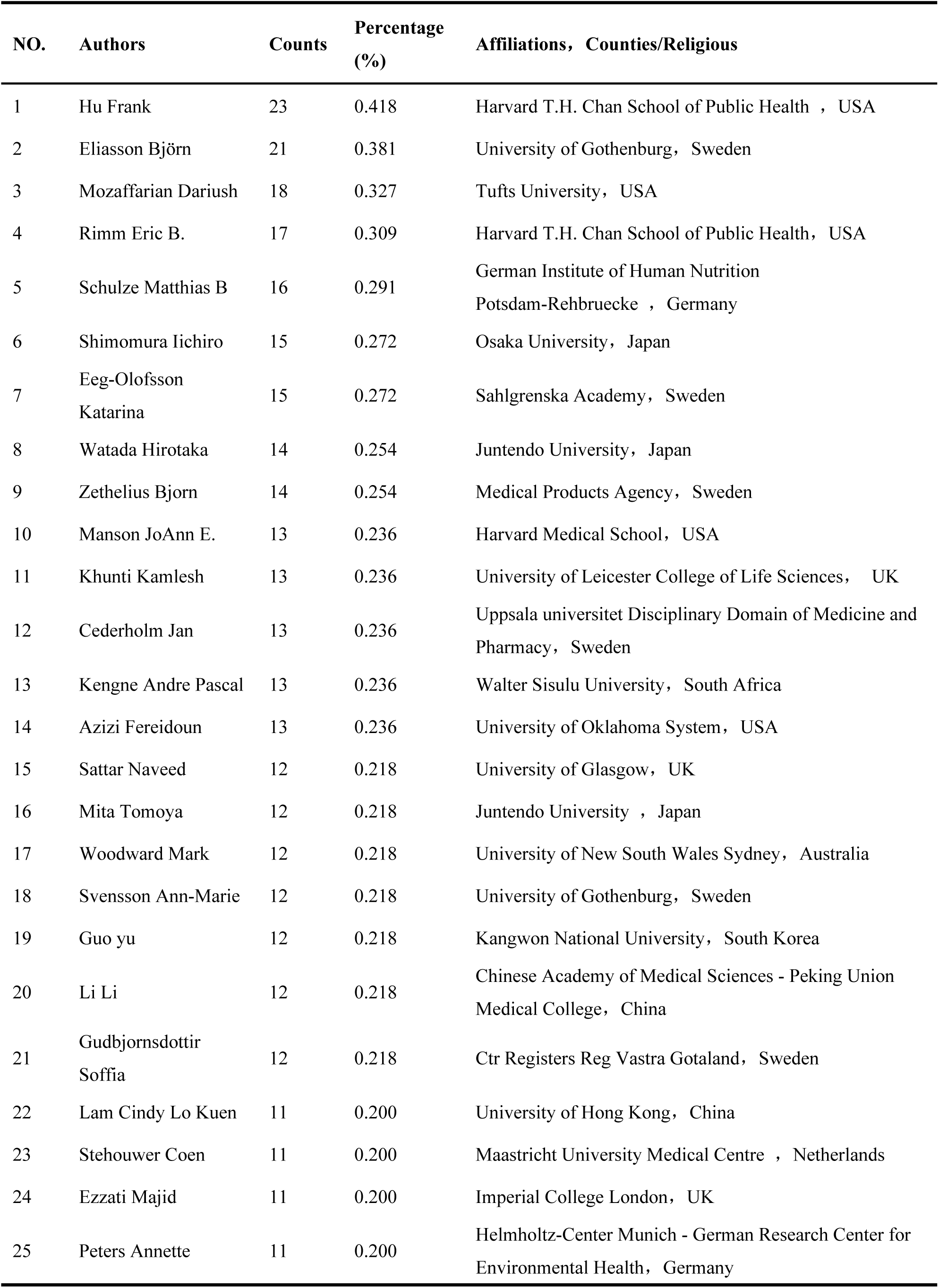
Top 25 authors in the filed of CVD in Diabetes from 2005 to 2024.

**TABLE 3.**
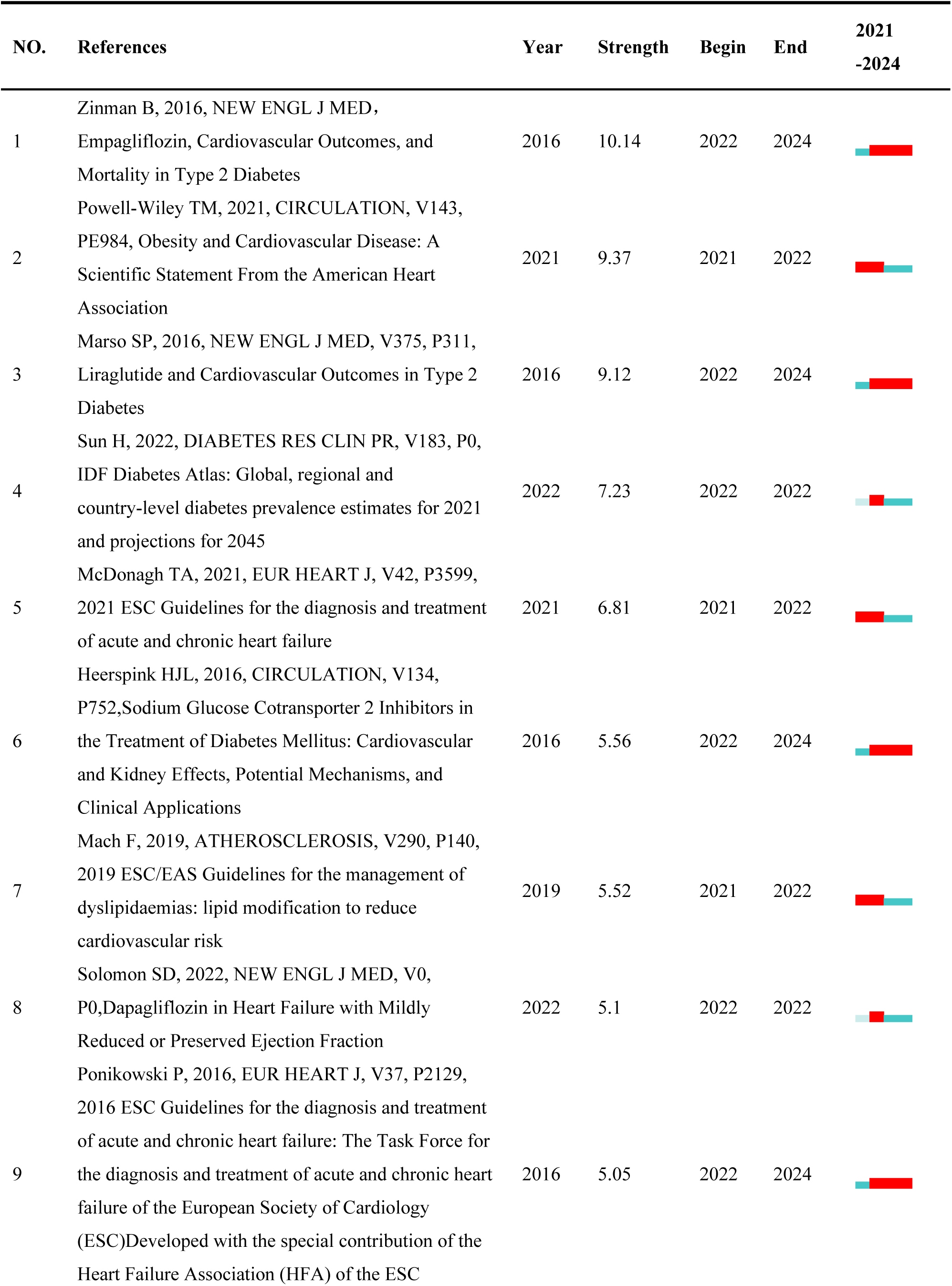
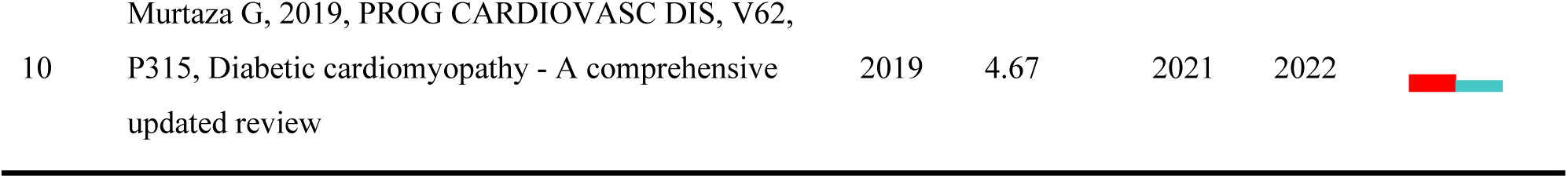
Top 10 references with the strongest citation bursts in the DM+CVD field (2021-2024)

### 2.2 Collaboration Network Analysis

From 2005 to 2024, research in the DM+CVD field involved contributions from 159 countries/regions, 7,137 institutions, and 34,707 authors. As shown in Figure 2, international collaboration in this field exhibits distinct geographical distribution patterns. Figure 2(A) displays the top 25 countries ranked by collaboration intensity, with European countries being particularly densely represented, indicating active research cooperation in this region. Regarding inter-country collaborations, the connecting lines between the United States and China, as well as between the United States and the United Kingdom, are particularly thick, reflecting close collaborative relationships among these countries.Figure 2(B) presents the top 10 countries/regions ranked by publication output. The United States leads with 1,257 publications, while China ranks second with 1,032 publications. Notably, there is a significant gap in publication numbers between the United Kingdom (ranked third) and subsequent countries compared to the United States and China.A comprehensive analysis of collaboration networks and publication data highlights the prominent positions of the United States and China in this research field, demonstrating their global academic influence and research capabilities.

**FIG 2.**
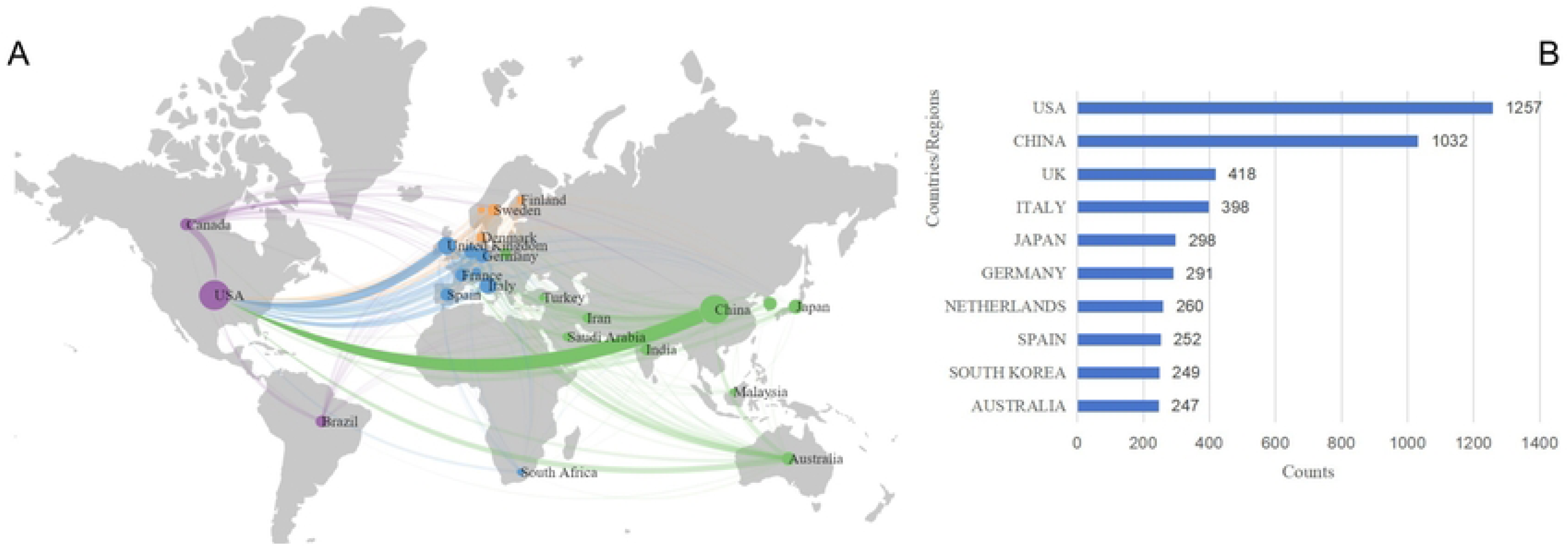
Map of countries/regions network cooperation (A) and top 10 countries/regions(B) in DM+CVD. (A) World map showing international collaboration networks. Lines represent collaborative links between countries, with different colors indicating distinct collaboration clusters. (B) Bar chart of the top 10 countries by publication count. The United States (1287 publications) and China (1032 publications) lead in research output, followed by the United Kingdom, Italy, and Japan.

The institutional collaboration network is depicted in Figure 3. In Figure 3(A), node size represents each institution’s publication output, while Figure 3(B) displays specific publication data. Harvard University tops the list with 226 publications, followed by Harvard Medical School (139) and University of London (134). Notably, 60% of the top 10 institutions by publication count are from the USA, highlighting the country’s dominant position and substantial contributions in this research field. This phenomenon also reflects the research strength and innovative capacity of American academic institutions in this domain.

**FIG.3.**
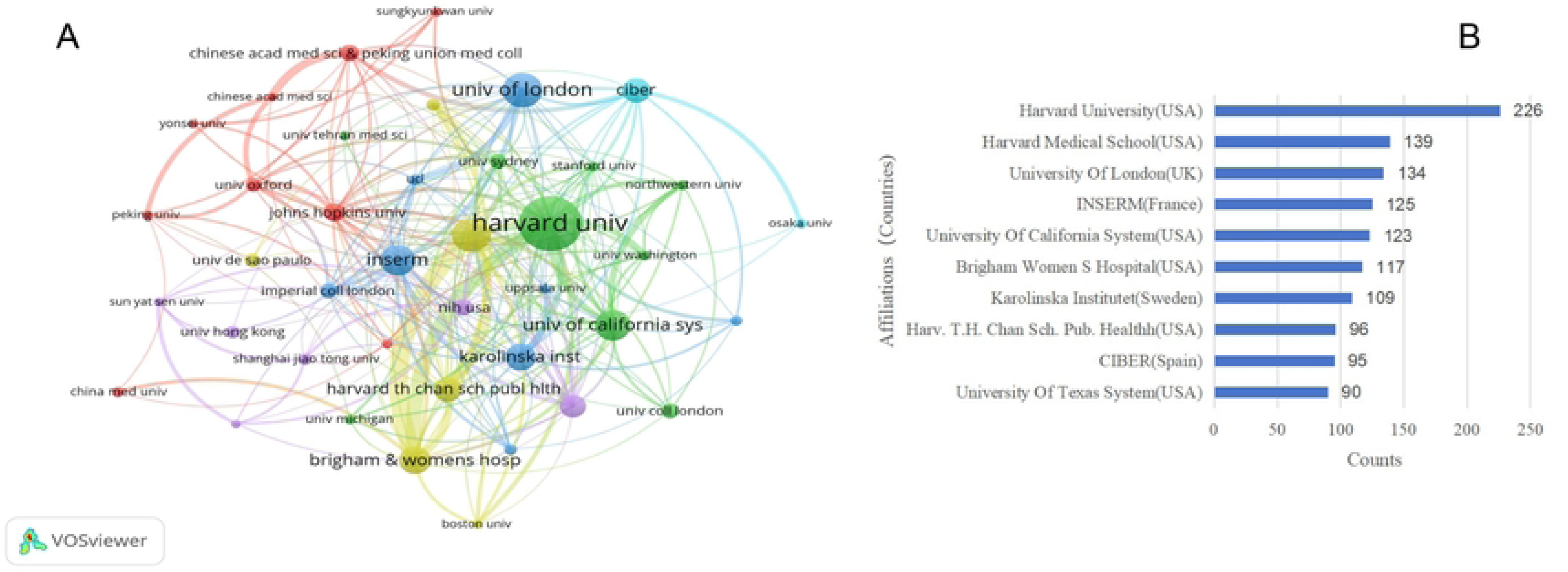
Institutional Cooperation Network (A) and top 10 Affiliations(B) in DM+CVD. (A) Visualization of institutional collaboration network. Nodes represent institutions, with size indicating publication volume. Colors denote different collaboration clusters. Lines represent collaborative links between institutions. (B) Bar chart showing top 10 institutions by publication count. Harvard University (USA) leads with 208 publications, followed by Harvard Medical School (USA) with 139 publications.

Figure 4 illustrates the author collaboration network and the top 10 authors by publication count.The intensity of the connecting lines in the network reflects the closeness of collaboration among authors. The top 10 authors typically have dense connections with other researchers, indicating their key roles in the collaborative network. It is noteworthy that some Chinese scholars, including Yang Ling, Chen Junshi, and Guo Yu, also show dense interconnections, reflecting the active engagement of Chinese research teams in this field.Among the top 25 ranked authors, 24% are Swedish, 20% are American, while among Asian countries, Japanese scholars comprise the highest proportion at 12%, followed by Chinese scholars at 8%. This distribution further accentuates the leading position of European and American countries in this field, while also signifying the increasingly significant roles of Asian countries, particularly Japan and China.

**FIG.4.**
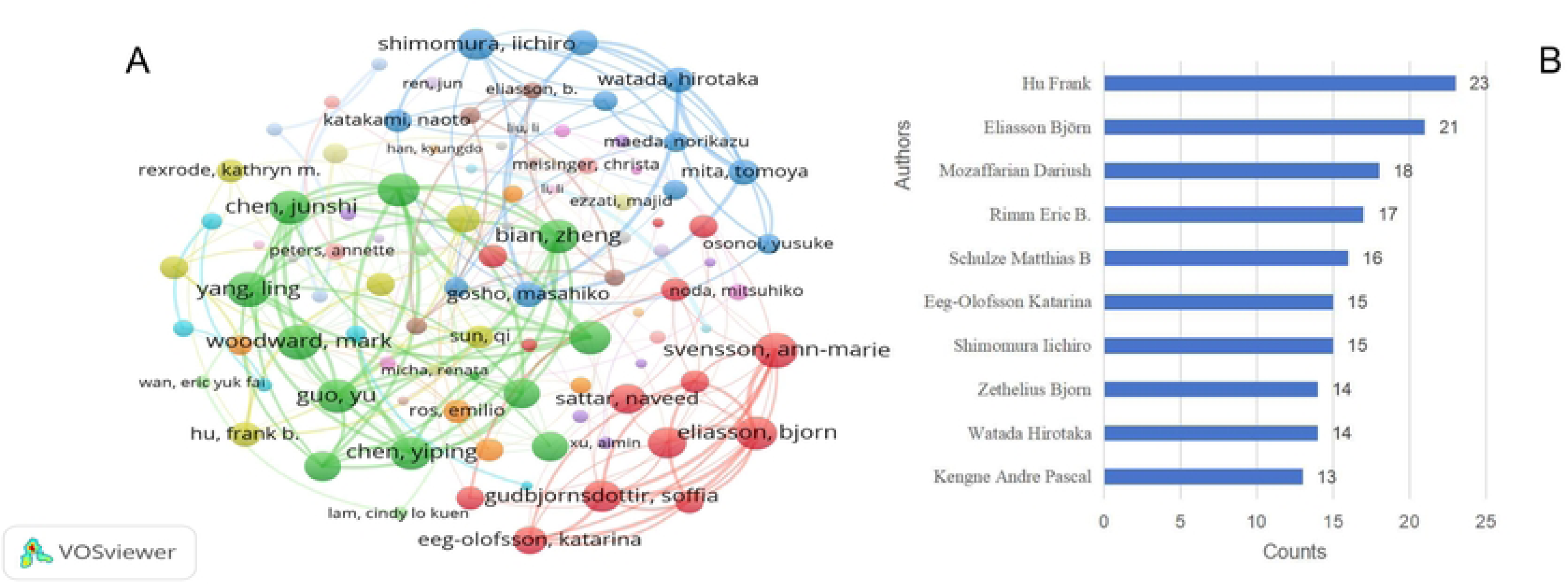
Author Cooperation Network (A) and top 10 Authors(B) in DM+CVD. (A) Visualization of author collaboration network. Nodes represent individual authors, with size indicating publication volume. Colors denote different collaboration clusters. Lines represent co-authorship links. (B) Bar chart showing top 10 authors by publication count.

In conclusion, within the interdisciplinary field of DM+CVD research, the United States, China, and European countries demonstrate notably high contributions. The United States maintains a leading position in overall research output, while European countries exhibit a more densely interconnected collaboration network. Asian countries, particularly China and Japan, also showcase robust research capabilities. This diverse landscape of international collaboration not only promotes knowledge exchange and innovation but also lays a solid foundation for the global development of this field.

### 2.3 Keyword Analysis: Dynamics and Trends

Through an analysis of keywords from 5,505 publications in the DM+CVD field, we systematically identified the research hotspots and developmental trends in this domain. VOSviewer data revealed a total of 8,250 keywords, with 16 keywords occurring more than 100 times. The keyword co-occurrence, cluster analysis, density distribution, and keyword bursts are illustrated in Figure 5.In the keyword co-occurrence network (Figure 5A), node size reflects keyword frequency. Larger nodes for terms such as “cardiovascular disease,” “diabetes mellitus,” and “obesity” align with the VOSviewer statistical data (Figures 5C, 5D) and correspond to our research focus. Other prominent keywords include “insulin resistance,” “hypertension,” “metabolic syndrome,” and “inflammation,” reflecting disease-related factors, risk factors, and pathophysiological processes associated with DM+CVD from various perspectives.Keywords like “gene expression” and “oxidative stress” indicate researchers’ interest in the fundamental mechanisms of DM+CVD. Terms such as “blood pressure” and “heart failure” highlight research hotspots in clinical manifestations and complications. Descriptive terms like “association” and “impact” correspond to scholars’ diverse research interests in different aspects of DM+CVD, reflecting to some extent the multidimensionality and complexity of research in this field.

**FIG 5.**
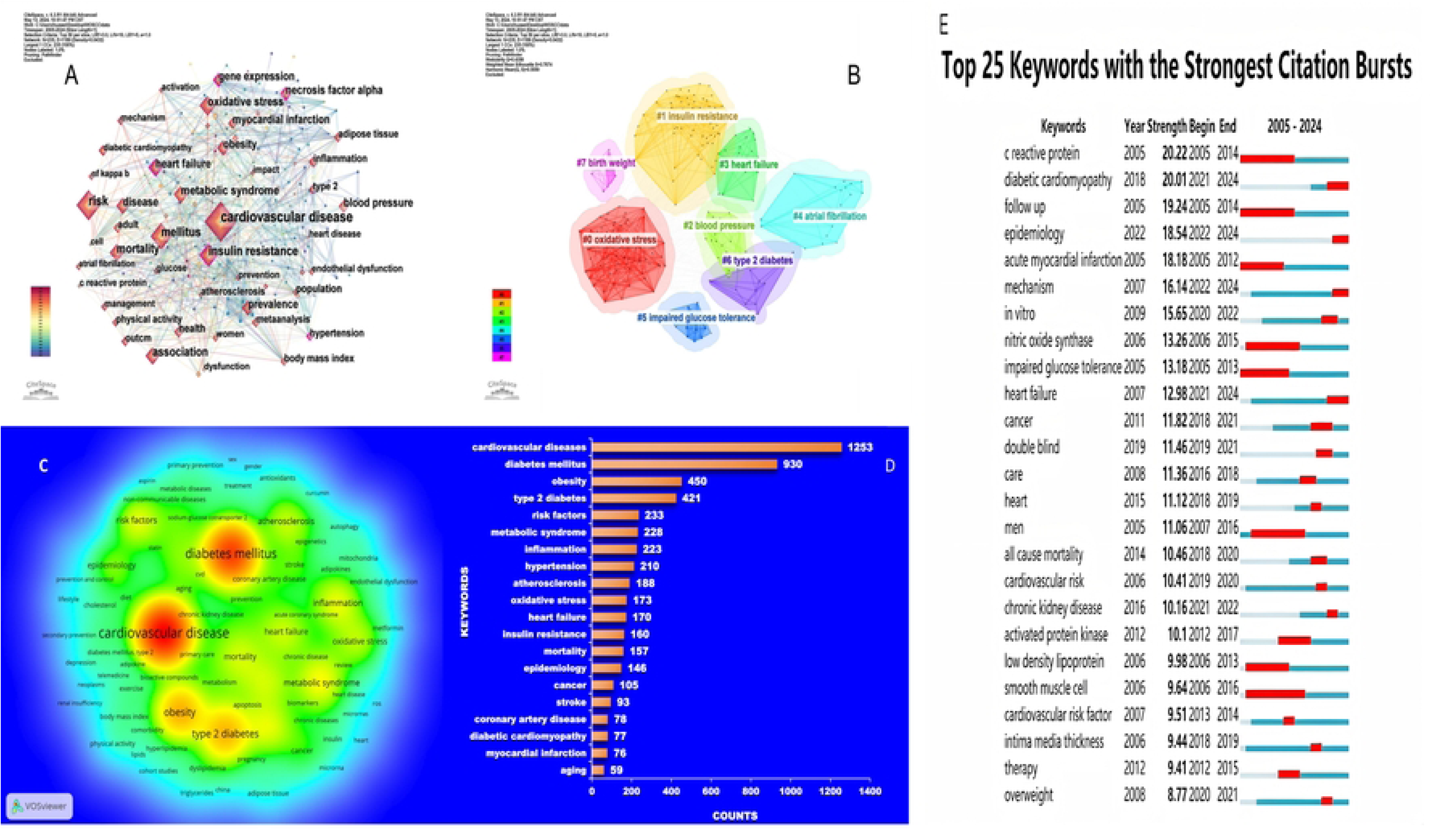
Keyword co-occurrence (A), clustering (B), density map (C), top 20 keywords (D) and Top 25keywords bursts(E) in the field of DM+CVD (2005-2024). (A) Keyword co-occurrence network. Node size indicates keyword frequency; colors represent different clusters.(B) Clusters of major research topics. Colored clusters show the evolution of research themes over time.(C) Density visualization of keyword co-occurrence. Red areas indicate high-frequency keywords and their relationships.(D) Bar chart of top 20 keywords by frequency. ‘Cardiovascular disease’ and ‘diabetes mellitus’ are the most frequent terms.(E) Top 25 keywords with the strongest citation bursts, showing emergence years and burst durations.

To better understand the underlying logic behind the keywords, we conducted a cluster analysis, with results shown in Figure 5B. The clustering indicators yielded a Q value of 0.4358 and an S value of 0.7674. Generally, Q > 0.3 indicates significantly effective clustering, S > 0.5 suggests reasonable clustering, and S > 0.7 denotes convincing clustering results17. The cluster analysis revealed the current research hotspots in DM+CVD:Clusters #0 (oxidative stress) and #1 (insulin resistance) indicate an in-depth exploration of fundamental pathological mechanisms. Clusters #2 (blood pressure), #3 (heart failure), and #4 (atrial fibrillation) reflect researchers’ emphasis on common clinical manifestations in DM+CVD patients. Clusters #5 (impaired glucose tolerance) and #6 (type 2 diabetes) demonstrate research interest in the entire disease progression process. The emergence of cluster #7 (birth weight) suggests that research has extended to early life stages, exploring factors that may influence the development of DM+CVD in adulthood.

Keyword burst analysis provides a crucial perspective for unveiling the dynamic evolution of the DM+CVD field. By identifying highly focused themes within specific periods, it offers insights into the shifts in research hotspots and helps capture emerging trends. Based on the keyword burst strength diagram generated by CiteSpace (Figure 5E), we conducted an in-depth analysis of the evolving research hotspots in the DM+CVD domain.The results indicate that C-reactive protein (CRP) exhibited the strongest burst strength (20.22) between 2005 and 2014, likely reflecting the intense research interest in inflammatory markers’ role in DM+CVD during this period. This was closely followed by diabetic cardiomyopathy (20.01, 2021-2024) and follow-up (19.24, 2005-2014). Notably, diabetic cardiomyopathy, as a relatively new research hotspot, has a burst period overlapping with the current timeframe, suggesting it may be one of the focal points of recent studies.Further analysis revealed multiple keywords showing burst trends in the 2021-2024 period, including the aforementioned diabetic cardiomyopathy, epidemiology (18.54), mechanism (16.14), and heart failure (12.98). These concurrent bursts may reflect the latest trends in DM+CVD research, namely an in-depth exploration of disease mechanisms, reassessment of epidemiological characteristics, and a focus on specific complications such as heart failure and diabetic cardiomyopathy.Additionally, we observed that certain keywords, such as “mechanism,” although present in research as early as 2007, did not show a significant burst trend until 2022. This lag phenomenon may indicate a shift in research focus from phenomenon description to mechanism exploration, reflecting an increase in knowledge accumulation and research depth within the field.Simultaneously, we noted that some earlier hotspots, such as CRP and acute myocardial infarction (18.18, 2005-2012), have concluded their burst periods. This may suggest that related research has matured or that research priorities have shifted.

The analysis of keywords and their evolutionary trends reveals the diversity and progression of research hotspots in the field. There has been a notable shift from the early focus on inflammatory markers and acute cardiovascular events to a more in-depth exploration of specific complications, such as diabetic cardiomyopathy and heart failure. The recently emerging keywords indicate that current research is concentrating on a deeper understanding of disease mechanisms and a reassessment of epidemiological characteristics. This progression demonstrates the continuous advancement of scientific knowledge in the DM+CVD field.

## 3. Analysis of Highly Cited Papers

Building upon our earlier keyword analysis that revealed research hotspots and trends in the DM+CVD field, we now turn our attention to the analysis of highly cited papers. These publications encapsulate the most significant scientific achievements in a discipline and represent the developmental level of the field. Analyzing highly cited papers allows for a comprehensive understanding of the structural framework and overall characteristics of the discipline.Figure 6 presents a dual-map overlay of research journals in the DM+CVD field, broadly illustrating two primary citation pathways. Published articles primarily fall within the domains of medicine, clinical research, molecular biology, and immunology, while cited literature predominantly originates from journals in health, nursing, medicine, molecular biology, and genomics. This reflects the interdisciplinary nature of research in this field and the balance between basic research and clinical applications.VOSviewer data indicate that 5,505 articles encompass 269,272 citations. As shown in Figure 7, there are 10 main clusters, including: #0 coronary disease, #1 treatment outcome, #2 heart failure, #3 myocardial infarction, #4 diabetes mellitus, #5 asymmetric dimethylarginine, #6 resveratrol, #7 epidemiology, #8 insulin resistance, #9 protein kinases, and #10 myocardial biology. These clusters reveal the core issues and research hotspots in the field.Through statistical and visual analysis of highly cited papers, combined with the citation report function of WOSCC, we discussed novel findings and key literature driving the development of the discipline, encompassing 28 articles and 22 reviews.

**FIG 6.**
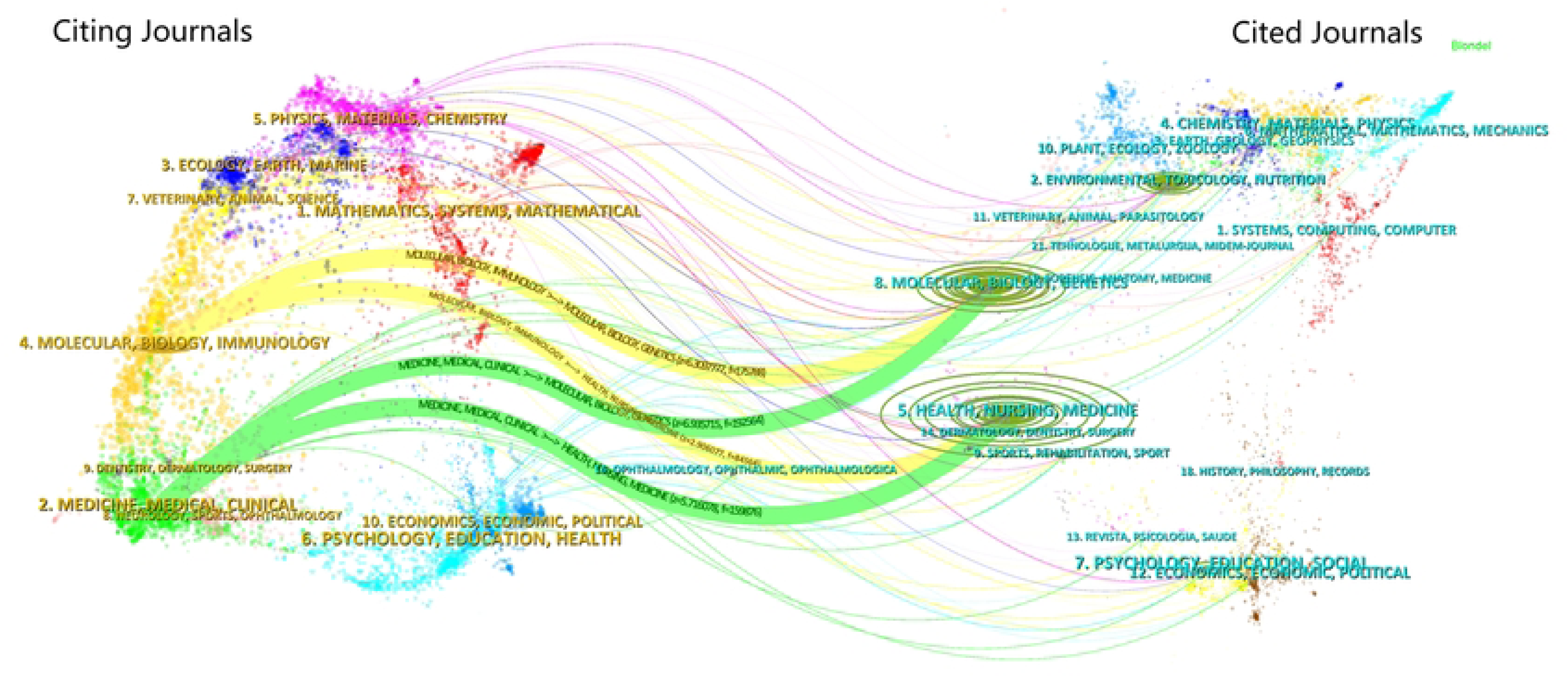
Dual-map overlay of the journals on DM+CVD research (2005-2024).Left side represents citing journals, right side represents cited journals. Each colored cluster represents a distinct research field. Lines connect citing journals to cited journals, showing interdisciplinary knowledge flow. Thickness of lines indicates citation frequency.

**FIG 7.**
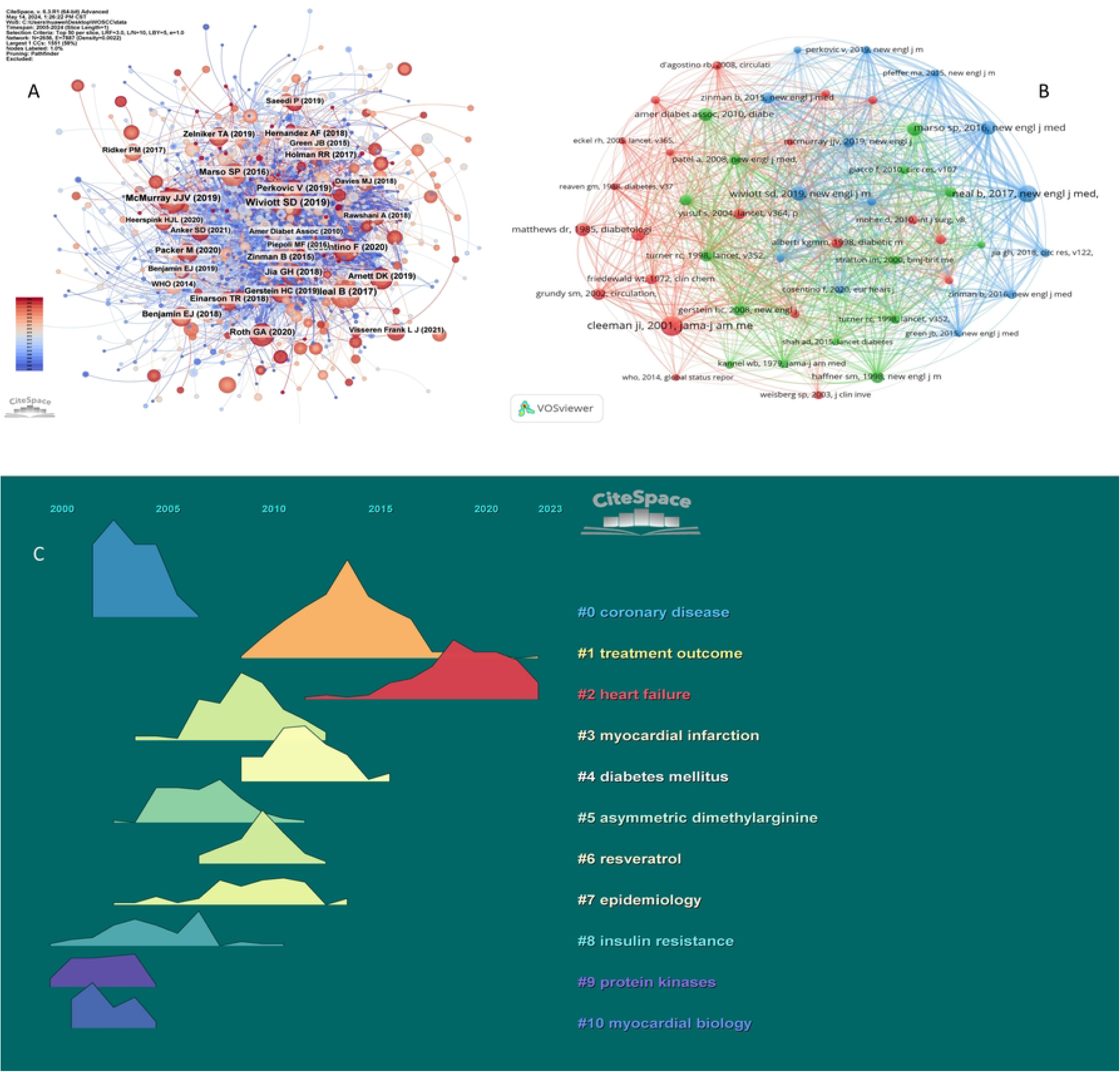
Co-citation network (A), top50 co-cited literature (B), and Timeline-based Theme River diagram (C) in DM+CVD(2005-2024). (A) Co-citation network of authors generated by CiteSpace. Node size represents citation frequency; colors indicate publication year (blue for older, red for more recent). (B) Co-citation network of literature generated by VOSviewer. Nodes represent publications; links indicate co-citation relationships. Different colors represent distinct clusters, each corresponding to a specific research theme or sub-field. (C) Landscape view of research topics from 2000 to 2023 created by CiteSpace. Each colored stream represents a major research theme, with labels indicating specific topics. The width of streams indicates the relative prominence of topics over time

### 3.1 Review

Reviews not only help researchers gain a comprehensive understanding of existing research findings and developmental trends but also facilitate knowledge accumulation and dissemination while identifying and addressing key issues through expert perspectives. To ensure research quality, authority, and impact, we selected 22 highly cited reviews published in top-tier journals such as The Lancet, Circulation Research, Circulation, Endocrine Reviews, European Heart Journal, and Cell Metabolism for analysis. These reviews have been categorized by research topic and ordered chronologically to distill the core information of their content.

#### Research on the Relationship Between Obesity and Disease

In 2005, Daniels SR et al.^19^reviewed the pathophysiological mechanisms of obesity in children and adolescents, elucidating how obesity leads to the development of DM band CVD through insulin resistance, chronic inflammation, and metabolic syndrome. This study not only provided crucial insights into understanding the pathogenesis of CVD in DM patients but also proposed multi-level prevention strategies encompassing lifestyle, family, community, and policy interventions, laying a theoretical foundation for subsequent research and policy formulation.In 2009, Daphne et al.^20^ conducted a meta-analysis synthesizing data from 89 studies. Their findings demonstrated a significant correlation between overweight/obesity and the risk of various chronic non-communicable diseases, particularly T2DM and CVD. By integrating extensive epidemiological data, this study provided robust evidence for understanding the association between overweight/obesity and DM/CVD.Prasenjit and Sushil^21^, in 2015, systematically explained how obesity exacerbates inflammatory responses through oxidative stress, thereby increasing the risk of T2DM and CVD. This deepened the understanding of the molecular mechanisms underlying obesity-related metabolic disorders.

As research progressed, Piché et al.^22^ proposed in 2020 that the phenotypic heterogeneity of obesity has diverse health impacts, with visceral fat accumulation significantly increasing the risk of CVD and DM. Further studies indicated that while overall obesity increases risk, the accumulation of fat in specific regions has a more pronounced effect on metabolic and cardiovascular health^23^.In 2021, the AHA issued a statement^24^ systematically summarizing the relationship between obesity and CVD. It explicitly identified obesity as an independent risk factor for CVD while emphasizing the close relationship between obesity and T2DM. Moreover, the statement highlighted key areas for future research, such as developing upstream interventions for young, severely obese patients for primary prevention and better chronic disease management. For older populations at risk of heart failure, strategies should focus on weight maintenance and improving functional outcomes rather than weight loss alone.

#### Public Health Strategies for Diet and Exercise

In 2010, Micha Renata et al.^25^ evaluated the differential health impacts of red and processed meat consumption. Their study indicated a significant positive correlation between processed meat intake and the risk of coronary heart disease and DM, while unprocessed red meat showed no such significant association. Subsequent research corroborated these findings, such as a meta-analysis published in *BMC Public Health*^26^ in 2019 and a study based on UK Biobank data^27^ in 2022. This accumulating scientific evidence has begun to influence global public health policies, with the World Health Organization^28^, the International Agency for Research on Cancer (IARC)^29^, and the European Society of Cardiology(ESC)^30^ all emphasizing the importance of reducing processed meat consumption.Beyond studies on specific foods, broader lifestyle interventions have also made significant progress. In 2015, Pedersen^31^ comprehensively reviewed the evidence for exercise as a treatment modality in 26 chronic diseases, highlighting the substantial effect of regular physical activity combined with dietary interventions in reducing DM and cardiovascular disease (CVD) risk. In 2016, Mozaffarian Dariush^32^ published a review in *Circulation*, integrating the latest dietary scientific evidence and emphasizing the importance of specific foods and overall dietary patterns (such as the Mediterranean diet) in preventing and managing chronic diseases.

Meanwhile, polyphenol research has provided new perspectives on lifestyle interventions. A 2015 review^33^ indicated that antioxidant phytochemicals like polyphenols can effectively scavenge excess oxidants, reducing oxidative stress levels. In 2018, Nour et al.^34^ further elucidated the immunomodulatory and anti-inflammatory mechanisms of polyphenols, including immune cell regulation, inhibition of pro-inflammatory factors, and inactivation of NF-kB. These mechanisms demonstrate the potential benefits of polyphenols in preventing and managing various chronic diseases, particularly in improving insulin sensitivity and vascular function. In 2020, Mehdi et al.^35^ conducted an in-depth analysis of the relationship between oxidative stress and various metabolic diseases, proposing lifestyle adjustments (such as increasing antioxidant intake and maintaining regular exercise) to improve oxidative stress status and thereby reduce disease burden. This research further reinforced the importance of antioxidants like polyphenols in chronic disease prevention.

However, despite encouraging results from polyphenol research, Hannah et al.’s review^36^ pointed out that currently only a few specific polyphenol compounds have received official approval for health claims, highlighting the need for enhanced regulation and research in this area. Additionally, the impact of food processing on polyphenol bioavailability requires further investigation to optimize their application in daily diets.

#### Theoretical Advances in the Mechanistic Research of DM+CVD

The pathophysiological mechanism research of DM+CVD involves complex interactions across multiple levels and disciplines. Through systematic analysis of highly cited literature, we have identified three main research directions: (1) Reassessmentof adipose tissue function, (2) In-depth study of cellular signaling pathways, and (3) Regulatory role of miRNAs in disease progressionThese studies reveal the complex pathophysiological processes of the disease from different perspectives, providing important clues for understanding disease mechanisms and developing new therapeutic strategies.

The reassessment of adipose tissue function is an important breakthrough in DM+CVD research. In 2008, Hajer et al. published a comprehensive review in the European Heart Journal^37^, summarizing the significant advances in the scientific community’s understanding of adipose tissue function at that time. This review emphasized that adipose tissue is not merely a simple fat storage site, but plays a key role in metabolic regulation and endocrine function.Adipose tissue directly participates in the pathophysiological processes of DM and CVD by secreting various hormones and cytokines, such as tumor necrosis factor-α (TNF-α), interleukin-6 (IL-6), and plasminogen activator inhibitor-1 (PAI-1). These findings provided new perspectives for subsequent research, prompting researchers to reconsider the associations between obesity, DM, and CVD

Research on cellular signaling pathways has further deepened our understanding of disease mechanisms. In 2010, Geraldes Pedro and King George L^38^ reviewed the potential role of protein kinase C (PKC) isoforms in the treatment of DM complications, which garnered widespread attention in the research community^39^. Although Ruboxistaurin, a PKC-β inhibitor, has shown promise in the treatment of diabetic retinopathy, further studies are needed to confirm its efficacy and safety in cardiovascular protection^40^.

Research on microRNAs (miRNAs) has provided new insights into disease mechanisms^41,42^. In 2012, Creemers Esther E et al.^43^ reviewed the progress in circulating miRNAs (miRNAs present in body fluids such as blood) research in CVD, highlighting their potential value as biomarkers. While the potential of circulating miRNAs as biomarkers has gained widespread attention, the fundamental challenges in their clinical application may be more profound. These include large individual variations in circulating miRNA expression^44^ standardization issues in detection methods^45^, consistency in sample collection and processing^46^ and the complexity in interpreting the origins and functions of circulating miRNAs^47^. Currently, these challenges limit the reliability of circulating miRNAs as diagnostic tools.

#### In terms of therapeutic theory research

Recent studies have not only deepened our understanding of existing treatments but also paved the way for personalized and precision medicine.Metformin, a cornerstone in DM treatment, has been a focus of research regarding its mechanism of action and clinical applications. A highly cited paper published in *Cell Metabolism* in 2014^48^elaborated on its mechanism and potential cardiovascular benefits. Despite metformin’s crucial role in DM treatment, its use may increase the risk of lactic acidosis in patients with renal impairment and high-risk cardiovascular conditions^49^, necessitating caution. These challenges, however, present opportunities for precision medicine, enabling more precise and safer individualized treatment strategies through the study of drug characteristics in different populations and clinical contexts.In 2016, Wang et al.^50^ comprehensively updated recent research findings in a review article in Circulation. They noted that current treatment strategies primarily focus on glycemic control, overlooking other important mechanisms by which DM exacerbates atherosclerosis and heart failure, such as chronic inflammation and endothelial dysfunction, which may vary in presentation and impact among patients. These findings highlight the necessity of personalized treatment approaches^51^.In the same year, Kautzky-Willer et al.^52^ further emphasized the importance of individualized treatment by analyzing the multifaceted impact of gender differences in T2DM. This study provided crucial evidence for developing gender-specific treatment strategies, though more clinical trials are needed to validate their efficacy.Forrester et al.^53^, in a 2018 *Circulation Research* paper, delved into the mechanisms of reactive oxygen species (ROS) in metabolic and inflammatory signaling, discussing potential strategies for treating related diseases by modulating ROS levels and signaling pathways. This study not only deepened the understanding of oxidative stress in DM+CVD but also suggested new approaches for targeting ROS-related pathways. However, translating these basic research findings into clinical applications faces numerous challenges, including precise control of ROS levels, potential dose-dependent side effects, and individual variations in ROS metabolism, all of which require further investigation. These challenges continue to drive the development of precision medicine in the field of DM+CVD.

### 3.2 Article

We analyzed 28 articles from the top 50 highly cited papers. Based on their research topics, we categorized these articles into three main groups: epidemiological studies, clinical research advancements in DM+CVD diagnosis and treatment, and management strategies.

#### In terms of epidemiological research

we conducted a comprehensive analysis of 7 relevant highly cited publications. We integrated the associated data and perspectives, summarizing their content and significant implications for public health.

he incidence and prevalence of CVD and DM continue to rise, and despite an overall slowdown in mortality rates, these two diseases remain among the leading causes of death globally^54,55^ The age-standardized prevalence of DM worldwide increased by 90.5% from 1990 to 2021, with the number of patients rising from 108 million in 1980 to approximately 529 million in 2021, of which 96% were T2DM patients^56^.The risk of CVD prevalence increases with age. Data from the NHLBI’s Framingham Heart Study (FHS) show^57^ that between 1980 and 2003, the annual incidence of first CVD events rose from 3/1000 in men aged 35-44 to 74/1000 in men aged 85-94. The number of CVD cases increased from 271 million in 1990 to 523 million in 2019^58,59^. Regarding mortality, the age-standardized death rate (per 100,000 population) for DM increased from 18.2 in 1990 to 19.6 in 2021. Although the age-standardized death rate for cardiovascular and circulatory system diseases decreased by 22% between 1990 and 2013^55^, considering global demographic changes, the number of CVD deaths continues to rise. As of 2021, CVD deaths still accounted for one-third of global deaths^59^.

There is a strong link between DM and CVD. Studies show^47^ that for every 10-year increase in DM duration, the risk-factor-adjusted CVD risk increases by 1.38-fold, and the mortality risk increases by 1.86-fold. Although the incidence of CVD events among adult DM patients has decreased by 50% since the beginning of the 21st century, the absolute risk of CVD remains twice that of non-DM populations^53^.DM+CVD imposes a significant disease burden on global public health, with notable regional disparities^54,55,60^ In economically developed countries such as the United States and Western European nations, despite declining mortality rates for DM and CVD, incidence continues to rise, primarily attributed to population aging and increasing obesity rates^57,61^. In contrast, low- and middle-income countries exhibit high rates of both incidence and mortality for DM and CVD, potentially due to lower education levels and awareness of prevention and control, weaker treatment capabilities, and insufficient healthcare resources^62–64^.

These studies collectively emphasize the persistent nature of DM and CVD as major global health threats. They provide crucial evidence for developing effective public health strategies to address current health challenges, particularly in areas such as resource allocation, implementation of preventive measures, and healthcare system reforms. Future research should further explore prevention and management strategies for these diseases, especially in resource-limited regions.

#### Clinical research advancements in DM+CVD diagnosis and treatment

Among the highly cited publications, 11 articles are related to this topic, covering biomarker studies, inflammation, and pharmaceutical research.

Biomarker researchplays a crucial role in the diagnosis and management of DM and CVD^65^, deepening our understanding of disease mechanisms and providing new directions for diagnostic, preventive, and therapeutic strategies. In 2007, Goralski et al.^66^ identified Chemerin as a novel adipokine that promotes insulin resistance by influencing insulin signaling and glucose metabolism, emerging as a new biomarker for DM^67^. Moreover, Chemerin increases CVD risk by promoting vascular inflammation and endothelial dysfunction, making it significant in cardiovascular risk assessment^68^.In 2017, Ellulu et al. provided a multidimensional perspective on DM+CVD biomarker research69, revealing the complex interactions between obesity, chronic inflammation, and oxidative stress in DM+CVD pathogenesis. They found a negative correlation between pro-inflammatory factors (TNF-α, IL-6) and adiponectin levels, while CRP levels strongly correlated with CVD risk. This finding not only linked metabolic disorders with vascular pathology but also provided a theoretical basis for multi-target therapeutic strategies.

However, these highly cited studies also reveal several challenges in biomarker application. Firstly, single biomarkers often fail to fully reflect the complex pathophysiological characteristics of DM and CVD^69^ suggesting that future research may need to develop multi-marker combination models. Secondly, these studies emphasize the importance of individual differences, as factors such as race, age, and gender may influence biomarker expression and significance^70^,.Overall, the rapid advancement in biomarker research has opened new opportunities for elucidating mechanisms, diagnosis, and treatment of DM+CVD. Nevertheless, to translate these research findings into clinical practice, more large-scale, multicenter prospective studies are needed to validate their clinical application value.

Inflammation plays a crucial role in understanding the pathophysiological mechanisms of diseases. In 2018, Ferrucci and Fabbri^71^ scientifically elucidated the roles of monocytes, macrophages, T cells, B cells, cellular senescence, and senescence-associated secretory phenotype (SASP) in CVD pathogenesis. These factors contribute to CVD risk by promoting the release of inflammatory mediators and exacerbating inflammatory responses. Numerous studies have corroborated this perspective^71–74^. Regulating endothelial nitric oxide synthase (eNOS) activity and function is an effective approach to mitigate inflammatory responses. However, eNOS “uncoupling” can transform it into a superoxide (O ₂ -) generating enzyme, thereby aggravating inflammation and CVD progression. Förstermann^75^ noted that increasing tetrahydrobiopterin (BH4) levels or inhibiting nicotinamide adenine dinucleotide phosphate (NADPH) oxidase activity could correct eNOS dysfunction, thus restoring its protective effects.

In the complex pathological network of DM+CVD, inflammatory response serves as a critical link, with implications far beyond mere cytokine release. DM patients, chronically exposed to hyperglycemia, not only experience low-grade inflammation but also undergo fundamental alterations in immune balance and metabolic regulation^76^. This systemic dysregulation accelerates atherosclerosis progression through multidimensional, multilayered mechanisms, creating a self-perpetuating cycle^77,78^.

Furthermore, we need to reconsider the dual role of inflammation in DM+CVD. While excessive inflammation is undoubtedly harmful, moderate inflammatory responses are necessary for tissue repair and metabolic homeostasis^79^. Therefore, future therapeutic strategies should aim not to completely eliminate inflammation, but rather to reestablish a healthy inflammatory balance, necessitating more refined regulatory approaches and long-term intervention strategies.

Pharmaceutical research. In recent years, DM and CVD-related drug research has shown trends towards diversification and precision. Analysis of highly cited literature reveals that hot topics in drug research focus on sodium-glucose cotransporter-2 inhibitors (SGLT2i), glucagon-like peptide-1 receptor agonists (GLP-1 RA), statins, antiplatelet drugs, renin-angiotensin-aldosterone system (RAAS) inhibitors, and β-blockers^80–82^. These drugs not only target glycemic control but also reflect multi-target intervention strategies for cardiovascular protection. The emergence of SGLT2i and GLP-1 RA, in particular, has provided new treatment options for DM+CVD patients, potentially altering clinical practice patterns.However, despite GLP-1 RA showing significant effects in DM treatment^83^, its role in preventing cardiovascular events remains controversial. A large-scale randomized controlled trial (RCT) by Pfeffer et al.^80^demonstrated that lixisenatide was non-inferior to placebo in terms of major cardiovascular endpoint incidence in T2DM patients with acute coronary syndrome, but failed to prove its superiority. This finding challenges our traditional understanding of incremental drug efficacy while highlighting the difficulty of evaluating single intervention effects in complex disease networks.

Another important point is the role of antiplatelet therapy in primary prevention of DM+CVD warrants further attention. Research by De Berardis et al.^84^revealed gender differences in the effects of low-dose aspirin and its potential bleeding risks. This finding not only emphasizes the importance of individualized treatment but also reflects the need for a more comprehensive consideration of the balance between risks and benefits when evaluating preventive interventions.

#### Comprehensive management strategies

Ten articles are related to this topic. The combined management of DM and CVD is a complex process aimed at comprehensively reducing cardiovascular risk and improving patients’ long-term prognosis and quality of life86. This requires consideration of multiple factors, including glycemic control, blood pressure management, lipid regulation (such as lowering LDL-C levels), weight management, and control of other cardiovascular risk factors. Achieving these goals necessitates addressing three core aspects: patient self-education and self-management, lifestyle interventions, and risk assessment.

Patient self-management and self-education have undergone significant evolution over the past two decades, progressing from basic disease awareness education to comprehensive multifactorial interventions, and now to technology-driven precision management. In 2007, the AHA and American Diabetes Association (ADA)^85^, along with the ESC and European Association for the Study of Diabetes (EASD)^86^, proposed fundamental frameworks for patient self-management in their respective joint guidelines, emphasizing disease awareness, lifestyle interventions, and medication adherence education. This phase focused on enhancing patients’ basic understanding of their conditions and encouraging proactive lifestyle changes.As medical knowledge deepened, the 2013 guidelines introduced a more comprehensive management concept^87^. This update expanded beyond singular glycemic control to integrate blood pressure and lipid management, while also incorporating psychological health into the overall management strategy.

From 2019 onwards^88–90^ guideline updates reflect characteristics of the digital age, ushering patient self-management into a new era. The introduction of smart devices and telemedicine has created unprecedented possibilities for real-time monitoring and precision management. For instance, the use of continuous glucose monitoring systems (CGM) and smart blood pressure monitors allows patients and healthcare teams to track key indicators more accurately. These technologies not only improve the precision and timeliness of data collection but also enable healthcare teams to respond more rapidly to patient needs, facilitating multidisciplinary collaboration and ongoing support.

Lifestyle interventions are an indispensable component in the comprehensive management of DM+CVD. Research by Danaei et al.^91^ corroborates the stance of major European and American guidelines, affirming that scientific lifestyle interventions, including dietary control, increased physical activity, and smoking cessation, not only help reduce CVD risk but also improve DM management outcomes. Weight management, as an integrated result of these interventions, emerges as a core objective in preventing and controlling both diseases.Mora et al.^92^further elucidated the importance of appropriate weight management, demonstrating that higher levels of physical activity can significantly reduce cardiovascular event risk through mechanisms affecting blood pressure and inflammation, among others. Concurrently, diet plays a crucial role in management strategies. Healthy dietary habits, particularly those rich in plant polyphenols^93^, offer cardiovascular protection by improving vascular function, lowering blood pressure, and reducing LDL oxidation.

While these studies provide evidence for the efficacy of lifestyle interventions, translating them into clinical practice remains challenging. Factors such as socioeconomic status, healthcare accessibility, and patient adherence often influence intervention outcomes^94^. Therefore, future efforts should focus on tailoring scientific interventions to patients’ social resources, sustainability, and individual characteristics. Additionally, exploring methods to enhance long-term adherence through digital health technologies warrants active investigation.

Risk assessment plays a crucial role in early disease prevention and individualized management. The Framingham Heart Study^95^ provided a multifactorial risk assessment model for evaluating overall and specific CVD risks in primary care. This multivariate risk assessment approach advanced cardiovascular disease prevention strategies and laid the foundation for subsequent risk assessment tool development. In recent years, risk assessment tools in the DM+CVD field have continually evolved, transitioning from traditional static models to more dynamic, personalized approaches. Updates to tools such as the QRISK3 cardiovascular risk assessment^96^, Finnish Diabetes Risk Score^97^, and dynamic heart failure risk prediction models^98^ showcase advantages in integrating more dynamic data sources, considering a broader range of risk factors, and utilizing advanced statistical and machine learning methods. These advancements enable clinicians to formulate more targeted prevention and treatment strategies. Regarding blood glucose and blood pressure monitoring, CGM and home blood pressure monitoring (HBPM) have become increasingly recommended methods. CGM technology provides real-time, continuous monitoring of blood glucose levels, offering DM patients valuable insights into glucose fluctuations^99^. HBPM supplies authentic blood pressure data from patients’ daily lives, avoiding interference from “white coat effect” and “masked hypertension”. HBPM data contributes to more accurate assessment of antihypertensive treatment efficacy, facilitating timely medication adjustments. Long-term HBPM data can also reveal circadian blood pressure rhythm changes, providing clues for identifying abnormal blood pressure patterns.

In conclusion, these studies indicate that management strategies for DM+CVD are evolving towards more integrated, precise, and personalized approaches. The development of technology-driven precision management models, individualized lifestyle intervention programs, and dynamic risk assessment tools presents increased possibilities for improving the efficacy of DM+CVD combined management. However, it is important to acknowledge that implementing these comprehensive management strategies still faces challenges in translating theory into practice, particularly when considering individual patient differences and socioeconomic factors. Future research should focus on enhancing the translational impact of new technologies, improving patient adherence, and developing more individualized and dynamic risk assessment models to achieve precision management of DM and CVD.

## 4. Research Hotspots and Development Trends

We attempted to discuss the research hotspots and trends in the field of DM+CVD by analyzing highly co-cited literature and their keywords from the past three years (2021-2023). The Citespace keyword analysis and its Spotlight function results are shown in Figure 8A. The Spotlight function of Citespace helps researchers quickly identify keywords that are widely cited and have significant impact on the current research field^17^. The results indicate that the main keywords are Cardiovascular disease, Risk, Oxidative stress, Inflammation, Obesity, Heart failure, Body mass index, Adipose tissue, Mortality, and Insulin resistance. This suggests that current research trends continue to revolve around these important and persistent hot topics.

**FIG 8.**
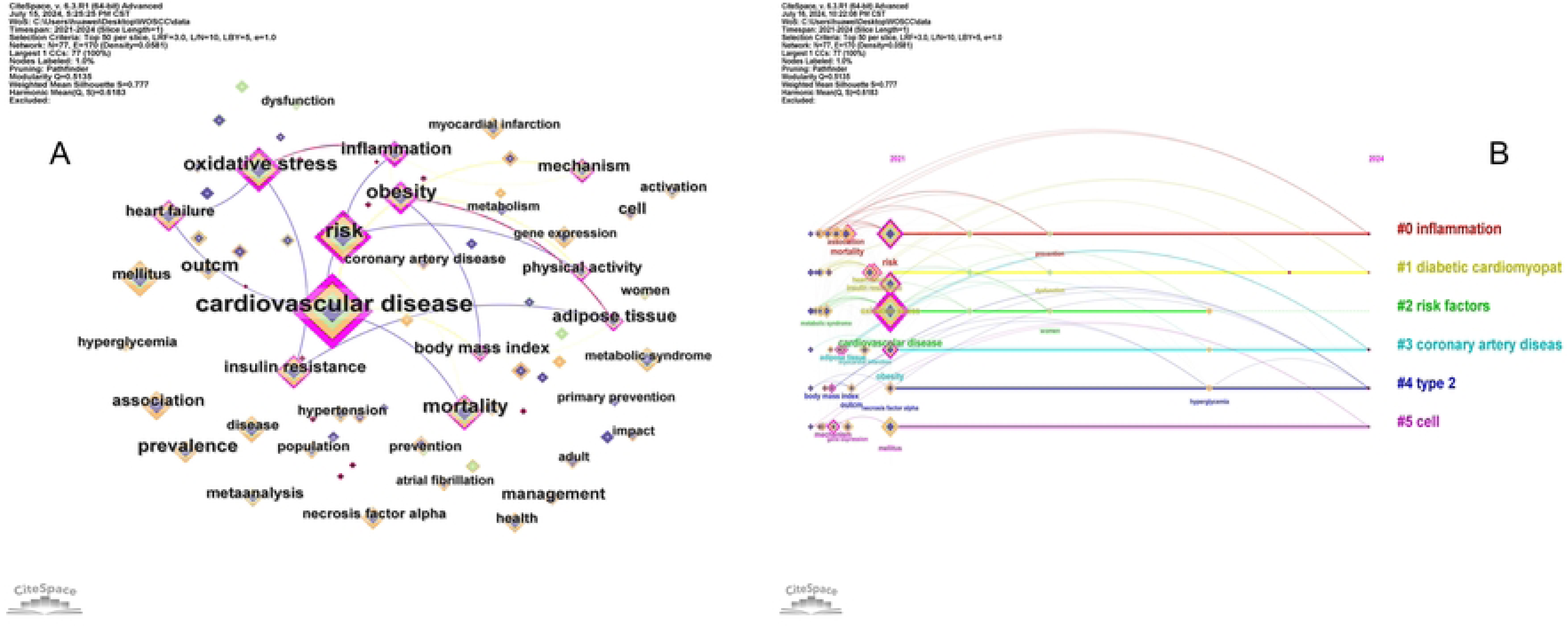
Visualization of Highly Cited Literature Keywords and Time line in DM+CVD (2021-2024) (A) Keyword co-occurrence network generated by CiteSpace. Node and font size represent the intensity and frequency of keyword occurrence. Node color indicates different time periods, with darker colors representing more recent times. Links between nodes, determined by the Spotlight algorithm, indicate high correlation between keywords. (B) Timeline view of major research topics. Each colored line represents a distinct research theme, with nodes on the lines indicating specific topics. The curvature and span of each line demonstrate the emergence and fading of different research themes over time.

Among the keywords, those in smaller fonts reflect emerging research directions gaining gradual attention, such as metabolic flexibility^100^, development of mitochondria-targeted therapeutic strategies^101^, and the role of epigenetics in cardiovascular diseases^102^. This indicates that researchers have been exploring more refined and individualized disease mechanisms and treatment approaches in recent years.Through the Spotlight algorithm, we observed significant associations between several groups of keywords, such as obesity and adipose tissue, cardiovascular disease and heart failure, and oxidative stress and inflammation. These close associations reflect the evolution and deepening of research focus over the past three years, while also revealing researchers’ in-depth exploration of internal connections within specific topics.Specifically, the link between obesity and adipose tissue indicates that research is delving into the complex relationship between obesity and adipose tissue function, particularly in terms of metabolic disorders and cardiovascular risk^103^. The association between cardiovascular disease and heart failure suggests the increasing importance of heart failure in DM+CVD research^104^, possibly related to population aging and improvements in chronic disease management. The close connection between oxidative stress and inflammation demonstrates researchers’ growing recognition of the interplay between these two processes in various pathological states^105^.

Clustering of keywords yielded six categories, as shown in Figure 8B: #0 Inflammation, #1 Diabetic cardiomyopathy, #2 Risk factors, #3 Coronary artery disease, #4 Type 2 diabetes, and #5 Cell. The Inflammation cluster emphasizes the central role of inflammation in DM+CVD pathology, possibly involving the development of novel anti-inflammatory therapeutic strategies. The Diabetic cardiomyopathy cluster suggests that researchers may be delving into the specific effects of DM on the myocardium. The Risk factors cluster reflects ongoing attention to DM+CVD risk prediction and prevention. The Coronary artery disease cluster underscores the pivotal position of coronary artery disease in DM+CVD. The Type 2 diabetes cluster emphasizes the close link between T2DM and CVD. The Cell cluster likely involves cellular-level mechanism studies, such as endothelial dysfunction or myocardial cell metabolic abnormalities.

Overall, these clusters span multiple levels from basic research to clinical applications, reflecting the complexity and multidisciplinary nature of DM+CVD research. They also hint at potential breakthroughs in future research. Future studies may focus more on the connections between these different themes to gain a more comprehensive understanding of DM+CVD.

In the previously mentioned trends in DM+CVD pharmacological research, SGLT2 inhibitors and GLP-1 receptor agonists have become the focus of recent studies due to their multiple clinical benefits. The proportion of highly cited literature in the past three years further confirms the importance of these drugs. Among the 10 articles, 4 involve SGLT2 inhibitors and GLP-1 receptor agonists.In 2016, Zinman et al^106^ published a study in *NEW ENGL J MED* with a median observation time of 3.1 years, concluding that for individuals with T2DM and high CVD risk, using SGLT2 inhibitors as adjunctive therapy reduced the rates of major cardiovascular events and all-cause mortality. In the same year, Marso et al^107^conducted a study with a median follow-up of 3.8 years, finding that liraglutide (a GLP-1 receptor agonist) significantly reduced cardiovascular event risk in T2DM, particularly cardiovascular death risk. This finding supports the inclusion of liraglutide in treatment regimens for type 2 diabetes patients with high cardiovascular risk.Also in 2016, Heerspink et al^81^ retrospectively examined the significant cardiovascular and renal protective effects of SGLT2 inhibitors in treating DM and discussed their clinical applications, suggesting that SGLT2 inhibitors should be prioritized when treating diabetic patients with high cardiovascular disease risk.In 2022, Solomon et al^108^conducted a randomized, double-blind, placebo-controlled trial, concluding that SGLT2 inhibitors significantly reduced the risk of worsening heart failure and cardiovascular death in patients with mildly reduced or preserved ejection fraction heart failure, while improving patient symptoms. The results of this study have significantly impacted clinical practice in using the SGLT2 inhibitor dapagliflozin to treat this type of heart failure patient.

Traditional T2DM management primarily focused on glycemic control, often utilizing insulin and sulfonylureas^109^. Cardiovascular disease prevention and treatment relied on statins, antihypertensives, and antiplatelet agents^110^. This segregated approach potentially led to several issues: dispersed treatment goals, increased risk of drug interactions, high medication burden for patients, potential side effects such as hypoglycemia and weight gain, lack of synergistic effects, and difficulties in achieving personalized precision treatment^109,111^.

In contrast, SGLT2 inhibitors and GLP-1 receptor agonists offer multifaceted clinical benefits in T2DM treatment. Moreover, these agents simultaneously improve glycemic control and cardiovascular health, thereby enhancing overall treatment efficacy and long-term prognosis^106–108^. Based on their comprehensive therapeutic effects and favorable safety profiles, authoritative bodies such as the ADA and EASD have prioritized SGLT2 inhibitors and GLP-1 receptor agonists in their clinical guidelines for specific high-risk diabetic patients^89,112^.Consequently, the mechanisms of action of SGLT2 inhibitors and GLP-1 receptor agonists remain a key focus of current and future research, particularly their physiological effects related to sodium metabolism. This ongoing investigation aims to better understand and apply these agents in the integrated management of DM and CVD.

Among the 10 articles, 2 discuss the association between lipids, obesity, and DM+CVD. The 2019 ESC/EAS^89^ guidelines on dyslipidemia management detail how to reduce CVD risk through lipid modulation. The guidelines emphasize early lipid risk intervention and lifestyle changes to effectively lower cardiovascular risk. For obese and DM patients, they particularly stress comprehensive management strategies to improve metabolic status and cardiovascular health. In 2021, the AHA released a scientific statement^24^ on obesity and cardiovascular disease, identifying obesity as a major risk factor for various cardiovascular diseases. It directly increases cardiovascular disease incidence by inducing metabolic abnormalities such as dyslipidemia, hypertension, and T2DM. The AHA emphasizes the importance of lifestyle interventions and weight loss for obesity prevention and treatment.

These two authoritative statements have profoundly influenced academia, clinical practice, public health policy, and public health education, advancing the comprehensive management and prevention of obesity and DM+CVD.

Although the relationships between obesity, DM, and CVD have been extensively studied, we believe current and future research trends will continue to focus on these areas. This ongoing focus is driven by several factors: the rising global prevalence of obesity^113^, DM, and CVD^59,64^; the crucial role of lipid metabolism abnormalities in these diseases^90^; the incompletely elucidated complex pathophysiological mechanisms; increasing demands for personalized medicine^114,115^; the emergence of novel therapeutic approaches; and the necessity for multidisciplinary comprehensive management^116^. Through multidisciplinary collaboration and personalized strategies, the aim is to comprehensively enhance the prevention and management of obesity, DM, lipid metabolism disorders, and CVD.

In the highly cited literature, three articles focus on the management of heart failure and cardiomyopathy. The 2016 ESC guidelines for heart failure treatment emphasized the importance of cardiac rehabilitation and self-management education, establishing the foundation for recent heart failure management^117^ These guidelines established ACE inhibitors, β-blockers, and MRAs as standard treatment regimens, detailed the diagnostic steps for heart failure, including clinical assessment, imaging, and laboratory tests, and introduced BNP and NT-proBNP as diagnostic tools. Additionally, they described acute heart failure management strategies in detail.Building on the 2016 guidelines, the 2021 ESC guidelines^118^ incorporated new drugs such as SGLT2 inhibitors and ARNIs, refined ejection fraction classification, strengthened the use of BNP and NT-proBNP, and further emphasized personalized treatment based on patient-specific factors, including age and comorbidities.In 2019, Murtaza et al. systematically reviewed the research progress on diabetic cardiomyopathy (DCM)^119^ exploring the roles of metabolic abnormalities, oxidative stress, and inflammatory responses in DCM pathogenesis. They highlighted the potential of novel drugs, gene therapy, stem cell therapy, and lifestyle interventions in future treatments. This comprehensive review significantly impacted the advancement of early diagnosis and effective treatment of DCM.

Although coronary heart disease remains a primary research focus, its study is relatively mature, with declining incidence and mortality rates in many countries and regions globally^59^. However, heart failure’s poor end-stage outcomes^120^ new opportunities for drug development^121–123^, prospects in regenerative medicine^124^ and the multiple complications and substantial medical burden of heart failure patients^125,126^ further underscore the importance and urgency of in-depth research into this disease.Moreover, the complex relationship between cardiomyopathy and diabetes, with their comorbidity and specific mechanisms not fully elucidated, adds another layer of complexity. The emergence of COVID-19 as a new etiological factor and the impact of long COVID have further complicated cardiomyopathy and increased its prevalence. Tegan et al.’s study^127^ found that COVID-19 patients have a 15.7-fold higher risk of myocarditis compared to non-infected individuals.

Considering these analyses, we believe that heart failure and cardiomyopathy will gradually become focal points of research in the DM+CVD field in the future.

Among the highly cited literature, one article focuses on epidemiological research. The International Diabetes Federation’s 2021 Diabetes Atlas (102nd edition)^128^ provides the latest data and projections on the global epidemiology of DM. This report reveals a significant growth trend in DM prevalence in developing countries and emphasizes the potential value of novel drugs such as SGLT2 inhibitors and GLP-1 receptor agonists in disease management. It also stresses the importance of early diagnosis and personalized treatment. Furthermore, the report proposes multifaceted intervention strategies, including strengthening public health education, promoting healthy lifestyles, and improving access to medical services and medications. This comprehensive and systematic report provides valuable reference for formulating DM prevention and control policies, holding significant implications for improving global DM prevention, treatment, and management.

Epidemiological research has consistently played a crucial role in DM+CVD prevention and control, and we believe it will remain a key research direction in the future^129^. Epidemiological studies not only help us understand disease distribution, development trends, and influencing factors but also provide scientific evidence for effective public health policy formulation^130^. Ongoing epidemiological surveillance and research are essential for evaluating the effectiveness of interventions, predicting disease burden, optimizing resource allocation, and developing long-term prevention and control strategies. Considering that DM patients often face a higher risk of CVD, epidemiological research has unique advantages in revealing the association between these two types of diseases, exploring common risk factors, and assessing the effectiveness of comprehensive prevention and control strategies^131^. In the future, with the development of big data and artificial intelligence technologies, epidemiological research is expected to play a greater role in disease warning, precision prevention, and personalized management, making more contributions to global DM and CVD prevention and control efforts.

In summary, we find that the current and future development directions in DM+CVD are not about replacing traditional pathways, but rather about posing new challenges and updates to existing treatment and research methods.Traditional sulfonylureas and statins have shown some efficacy in treating DM or CVD individually, but face challenges when addressing DM combined with CVD or more complex situations. The impact of obesity and abnormal lipid metabolism in DM+CVD patients cannot be overlooked; while weight reduction and lipid metabolism regulation have demonstrated significant clinical benefits in improving prognosis for these patients, their mechanisms of action and optimal intervention strategies are not yet fully elucidated.Although coronary heart disease still dominates DM+CVD research, heart failure and cardiomyopathy are gradually becoming focal points in this field, considering the various clinical scenarios in reality and the complexities associated with their poor end-stage manifestations. Epidemiological research is crucial for understanding the complex relationship between DM and CVD. As global population structure, socioeconomic environments, and risk factors change, such research helps identify and quantify risk factors, providing a scientific basis for developing prevention and management strategies.

Therefore, we summarize the current and future development directions as:

1. Intensifying research on novel drugs such as SGLT2 inhibitors and GLP-1 receptor agonists;
2. Updating our understanding of the key roles of obesity and lipid metabolism in disease progression;
3. Focusing on heart failure and cardiomyopathy as the main diseases of study in this field;
4. Strengthening epidemiological research on DM+CVD.

## 5. Conclusion and Future Perspectives

This study employs bibliometric methods, utilizing CiteSpace and VOSviewer analytical tools, to conduct a systematic analysis of 5,505 relevant publications from 2005 to 2024, aiming to elucidate research dynamics and future trends in the field. Initially, we examined and analyzed publication distribution, countries/regions, institutions, and author productivity and collaborations. We observed a significant growth trend in DM+CVD research, reflecting both sustained academic interest in the field and the increasing recognition of DM+CVD as a complex disease combination. Regarding research collaboration networks, we noted an interesting geographical distribution pattern. While the United States and China dominate in publication volume, European countries, particularly Sweden, stand out in rankings of highly productive authors and institutions. This distribution reflects the global nature of DM+CVD research while potentially indicating regional differences in research priorities.Subsequently, we focused on keywords and highly cited literature. After a comprehensive evaluation of the 5,505 publications from 2005-2024, we analyzed 50 highly cited papers in depth, including 22 reviews and 28 original research articles. Special attention was given to research developments in the past three years (2021-2023) to better understand future trends in the field.

The review analysis reveals the evolution of the theoretical framework in DM+CVD research. Specifically, in studying obesity and disease relationships, the perspective has shifted from viewing obesity as a simple disease risk factor to recognizing its phenotypic heterogeneity and differential impact on DM and CVD, ultimately emphasizing multi-level prevention and intervention strategies across the life course. In lifestyle intervention and public health research, the focus has progressed from examining the health effects of single food components to studying the combined role of overall dietary patterns and regular exercise, and finally to integrating dietary interventions into broader public health policy frameworks.Mechanistic studies have demonstrated in-depth exploration at multiple biological levels, including reassessment of tissue function, deeper investigation of cellular signaling pathways, and new discoveries in molecular regulatory mechanisms. Treatment strategies have advanced from single-disease management models to comprehensive risk control and individualized precision therapy.

Analysis of original research illustrates the development from theory to clinical practice. Epidemiological studies have quantified the global disease burden of DM+CVD, providing empirical evidence for evidence-based public health strategies. Clinical diagnosis and treatment research encompasses the identification of novel biomarkers, elucidation of inflammation’s central role in disease pathophysiology, and efficacy validation of new drugs. Management strategy research reflects a transition from basic awareness education to technology-driven, multi-factorial, and multi-disease comprehensive risk control.

Finally, through analysis of highly cited literature from the past three years, we identified or predicted future research priorities in the field, including: novel drug intervention strategies, pathophysiological associations between metabolic abnormalities and DM+CVD, precision management of cardiovascular complications, and more extensive epidemiological research.

In nearly two decades, despite significant advancements in this field, we have identified several persistent challenges. The long-term effects and precise mechanisms of novel drugs require further investigation. Clinical application of biomarkers faces issues of standardization and individualization. Achieving a precise balance in inflammation regulation remains a complex problem. Notably, effectively preventing and managing obesity at the population level to reduce DM+CVD incidence continues to be a major challenge.In general, DM+CVD research is evolving from descriptive and observational studies towards mechanism-oriented and personalized approaches, with obesity consistently emerging as a key factor. Looking ahead, interdisciplinary collaboration and translational medical research will play crucial roles in driving breakthrough progress in the DM+CVD field, particularly in deciphering the complex relationships between obesity and DM+CVD. This not only holds promise for improving patient outcomes but may also provide insights for managing other obesity-related complex chronic diseases.

## Data Availability

All relevant data are within the manuscript and its Supporting Information files.

## Acknowledgments

We thank the Science and Technology Department of Xinjiang Uygur Autonomous Region and the Science and Technology Bureau of Hetian Prefecture for supporting this research.

## Supplementary Table

**Table S1.** Thematic Areas for Meso-level Citation Analysis.

| NO. | Citation topics Meso |
| --- | --- |
| 1 | Diabetes |
| 2 | Phytochemicals |
| 3 | Lipids |
| 4 | Nutrition & Dietetics |
| 5 | Cardiology - Circulation |
| 6 | Cardiology - General |
| 7 | Healthcare Policy |
| 8 | Molecular & Cell Biology - Cancer |
| 9 | Autophagy & Apoptosis |
| 10 | Sleep Science & Circadian Systems |
| 11 | Molecular & Cell Biology - Pharmacology |
| 12 | Micro & Long Noncoding Rna |
| 13 | Obstetrics & Gynecology |
| 14 | Genome Studies |
| 15 | Immunology |
| 16 | Nursing |
| 17 | Psychiatry |
| 18 | Stem Cell Research |
| 19 | Molecular & Cell Biology - Mitochondria |
| 20 | Cardiac Arrhythmia |
| 21 | Strokes |
| 22 | Auto-inflammatory Diseases |
| 23 | Extracellular Matrix & Cell Differentiation |
| 24 | Vascular |
| 25 | Cardiac & Thoracic Surgery |
| 26 | Molecular & Cell Biology - Genetics |
| 27 | Molecular & Cell Biology - Physiology |
| 28 | Substance Abuse |
| 29 | Endocrinology & Metabolism |
| 30 | Smoking Cessation |
| 31 | Health Literacy & Telemedicine |
| 32 | Antibiotics & Antimicrobials |
| 33 | Social Psychology |
| 34 | Hematologic Diseases |

